## Supplementary figures and images for "Fatigue in Incident Peritoneal Dialysis and Mortality: A Real-World Side-by-Side Study in Brazil and the United States"

### S1 Fig

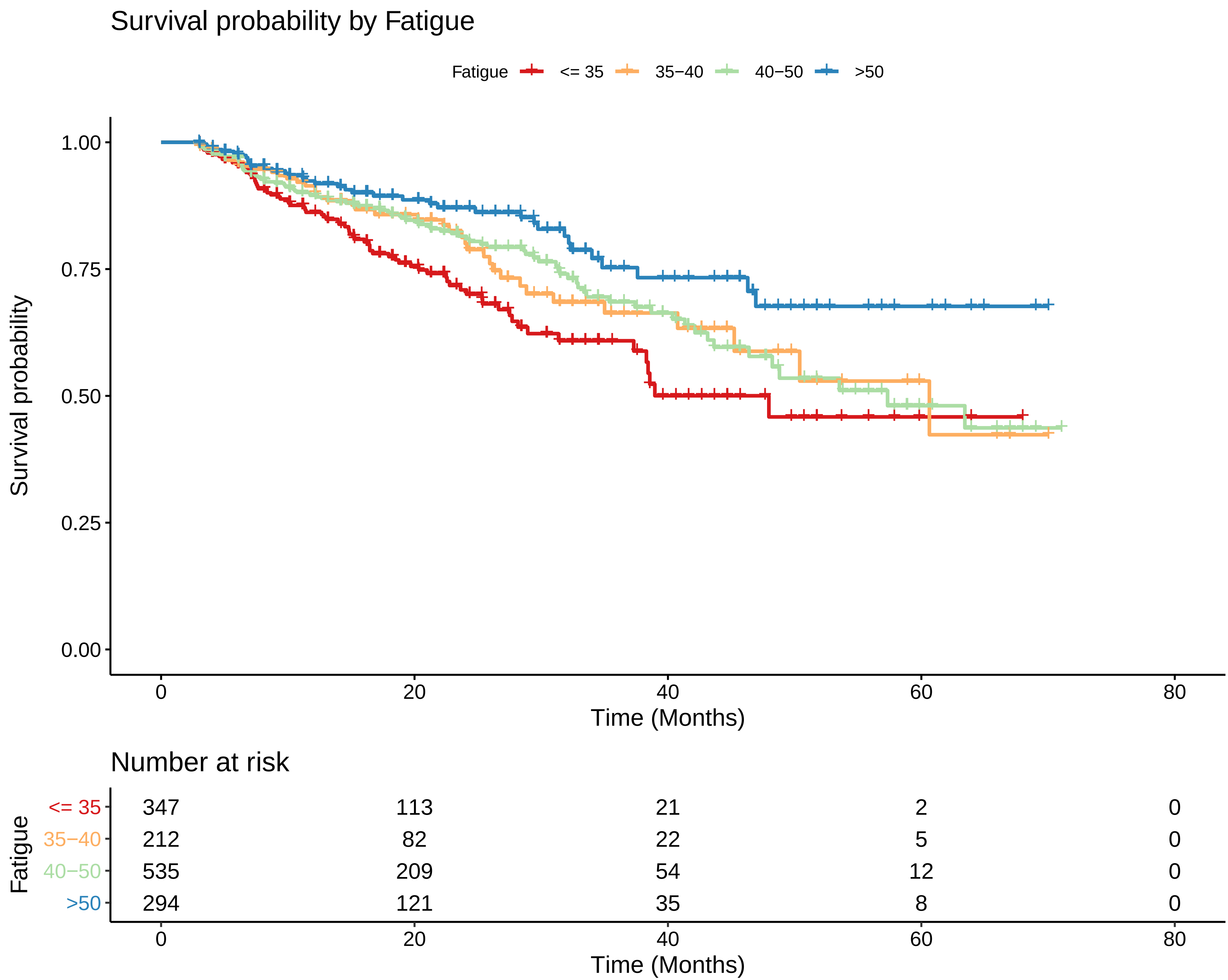

### S2 Fig

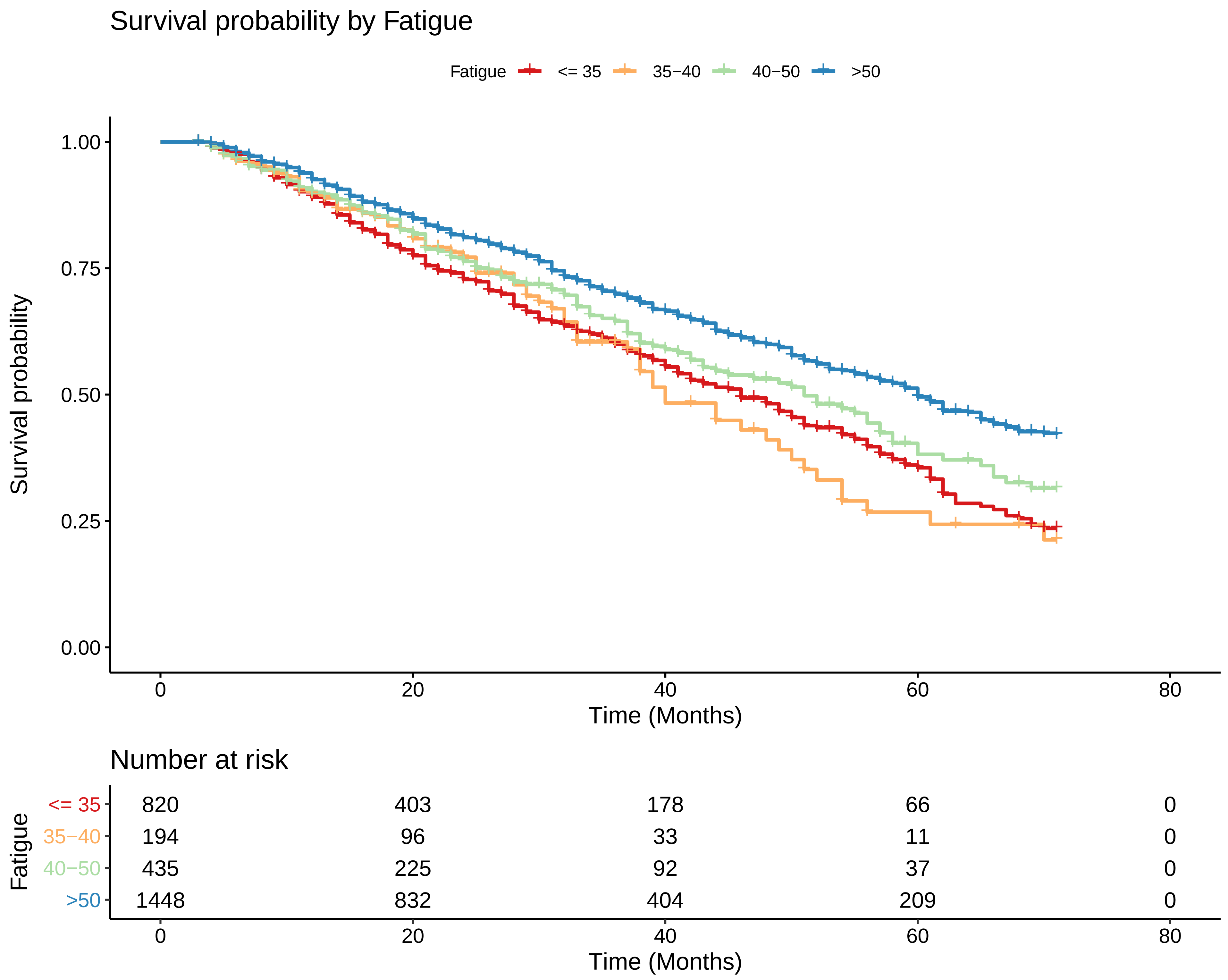
