## Supplementary material for "Fatigue in Incident Peritoneal Dialysis and Mortality: A Real-World Side-by-Side Study in Brazil and the United States": S1 Tab

| **Supplementary Table 1. Univariate model estimates for all-cause mortality risk in the Brazil and United States cohorts** | | | | | | |
| --- | --- | --- | --- | --- | --- | --- |
| **Cohort** | **Brazil Cohort** | | | **United States Cohort** | | |
|  | Estimate | Lower 95% CI | Upper 95% CI | Estimate | Lower 95% CI | Upper 95% CI |
| **Gender (Male)** | 1.09 | 0.86 | 1.4 | 1.11 | 0.98 | 1.27 |
| **Age > 65 years** | 2.54 | 1.98 | 3.26 | 2.34 | 2.06 | 2.67 |
| **Race (white)** | 1.22 | 0.94 | 1.57 | 1.72 | 1.47 | 2.01 |
| **Body mass index** | 0.98 | 0.95 | 1.01 | 1 | 0.98 | 1.01 |
| **Coronary artery disease** | 1.8 | 1.38 | 2.36 | 2.13 | 1.81 | 2.51 |
| **Diabetes** | 1.8 | 1.41 | 2.31 | 1.65 | 1.44 | 1.88 |
| **Initial Modality (CAPD)** | 0.78 | 0.6 | 1.01 | 1.06 | 0.93 | 1.2 |
| **Peripheral artery disease** | 2 | 1.54 | 2.58 | 2.08 | 1.68 | 2.59 |
| **Previous HD** | 1.25 | 0.98 | 1.6 | 1.52 | 1.34 | 1.73 |
| **Dialysis vintage (years)** | 0.96 | 0.94 | 0.97 | 1.01 | 1.01 | 1.01 |
| **Residual kidney function** | 0.64 | 0.5 | 0.82 | 0.88 | 0.54 | 1.46 |
| **Prescribed Volume 24h** | 1 | 1 | 1 | 1.1 | 1.04 | 1.15 |
| **Creatinine** | 0.92 | 0.88 | 0.97 | 0.96 | 0.94 | 0.98 |
| **Hgb** | 0.94 | 0.87 | 1.01 | 0.94 | 0.9 | 0.98 |
| Estimates present the hazard ratio and 95% confidence internals (CI) for all-cause mortality. CAPD: Continuous ambulatory peritoneal dialysis; HD: Hemodialysis. | | | | | | |
